# Plasma glucosylceramide levels are regulated by *ATP10D* and are not involved in Parkinson’s disease pathogenesis

**DOI:** 10.1101/2024.09.13.24313644

**Authors:** Emma N. Somerville, Alva James, Christian Beetz, Robert Schwieger, Gal Barrel, Krishna K. Kandaswamy, Marius I. Iurascu, Peter Bauer, Michael Ta, Hirotaka Iwaki, Konstantin Senkevich, Eric Yu, Roy N. Alcalay, Ziv Gan-Or

**Author notes:** **Corresponding author:** Ziv Gan-Or, Department of Neurology and Neurosurgery, McGill University, 1033 Pine Avenue West, Ludmer Pavilion, Room 312, Montréal, QC, Canada, H3A 1A1.

## Abstract

*GBA1* variants and decreased glucocerebrosidase (GCase) activity are implicated in Parkinson’s disease (PD). We investigated the hypothesis that increased levels of glucosylceramide (GlcCer), one of GCase main substrates, are involved in PD pathogenesis. Using multiple genetic methods, we show that *ATP10D*, not *GBA1*, is the main regulator of plasma GlcCer levels, yet it is not involved in PD pathogenesis. Plasma GlcCer levels were associated with PD, but not in a causative manner, and are not predictive of disease status. These results argue against targeting GlcCer in *GBA1*-PD and underscore the need to explore alternative mechanisms and biomarkers for PD.

## Introduction

Variants in *GBA1*, encoding the lysosomal hydrolase glucocerebrosidase (GCase), are common risk factors for Parkinson’s disease (PD). In most populations, *GBA1* variants have been identified in 5-20% of PD patients (*GBA1*-PD),^1^ and recently, a common intronic variant has been reported in ∼40% of African PD patients.^2^ *GBA1*-PD is characterized by earlier age at onset, higher frequency of non-motor symptoms, and rapid progression of both motor and non-motor symptoms.^3^ In addition to *GBA1*-PD patients, ∼1/3 of idiopathic PD patients who do not carry *GBA1* variants have reduced GCase activity compared to healthy controls.^4^ The reasons for this decreased activity are unknown, although several modifiers of GCase activity have been identified.^5-7^ Altogether, reduced GCase activity due to genetic variants or other reasons may be found in roughly half of PD patients.

Several hypotheses have been made on the pathogenic mechanism underlying *GBA1*-PD, and a central one suggests that increased levels of glucosylceramide (GlcCer, or GL-1), one of the main substrates of GCase, may lead to PD. This hypothesis states that GlcCer accumulates due to reduced GCase activity, and then interacts with alpha-synuclein leading to its accumulation.^8^ Based on this and subsequent results, a recent phase II trial examined the effect of venglustat, an inhibitor of GlcCer synthase, on PD progression (NCT02906020). The drug showed excellent target engagement, and indeed reduced the levels of GlcCer both in plasma and CSF. However, on average, there was no clinical benefit for the drug and in subanalyses carriers of the common p.N370S GBA1 variant who received the drug deteriorated faster than the placebo group.^9^ This observation argues against GL-1 levels being the mediator of the link between GBA-1 mutations and PD risk.

In this study, we applied multiple genetic methods to examine the association between GlcCer and PD, in order to examine whether there is genetic evidence for its involvement in the pathogenesis of PD.

## Methods

As a discovery cohort, we used the Parkinson’s Progression Markers Initiative (PPMI) population, including 366 PD cases, 164 controls, and 51 individuals with parkinsonism and scans without evidence of dopaminergic deficit (SWEDD, Supplementary Table 1). All subjects were of European descent, confirmed with principal component analysis (PCA). Informed consent forms were administered and signed by all participants before entering the study, and the study protocol was approved by the relevant institutional review boards. GlcCer in plasma was extracted by protein precipitation and analyzed with liquid chromatography-tandem mass spectrometry (LC-MS/MS) at Sanofi laboratories (Supplementary Text). Genotyping was performed on the NeuroX array, according to manufacturer’s protocols (Illumina Inc.). Quality control for individual and variant-level data was completed as previously described (https://github.com/neurogenetics/GWAS-pipeline). Imputation was performed with the TOPMed

Imputation Server using the TOPMed reference panel r3 and default settings.^10^ Genome-wide association studies (GWAS) were performed with linear regression in plink v1.9 using soft-called variants (R^2^ > 0.3) with a minor allele frequency (MAF) greater than 0.05.^11^ Adjustments for age, sex and the top 5 principal components (PCs) were included. PCs were calculated with PCA in plink v1.9.^11^ Conditional and joint analyses were performed with GTCA-COJO to identify independent variants after adjusting for lead SNPs.^12^ Linkage disequilibrium Manhattan plots were created using LocusZoom (http://locuszoom.org/). Regressions of GlcCer activity with PD status and area under the curve (AUC) analyses were performed R v4.3.2.^13^ Colocalization analysis was performed with coloc R package (https://chr1swallace.github.io/coloc/index.html) and plots were created with the LocusComparerR R package to plot GlcCer GWAS p-values against PD GWAS p-values (https://github.com/boxiangliu/locuscomparer). Linkage disequilibrium heatmap plots were generated using the LDmatrix tool through LDlink (https://ldlink.nih.gov/?tab=home).

Methodology for glucosylceramide extraction, genetic data processing, and analysis methods can be found in the Supplementary Text.

## Results

To identify common genetic variants associated with changes in plasma GlcCer levels, we performed a GWAS for each isoform. 489 individuals and 2,100,613 variants were included in each analysis after quality control and imputation, and mean levels for each isoform were calculated (Supplementary Table 2). Lambdas and QQ plots were assessed for all isoforms to test for systematic bias, and were deemed satisfactory (Supplementary Figure 1, Supplementary Table 3).

**Figure 1.**
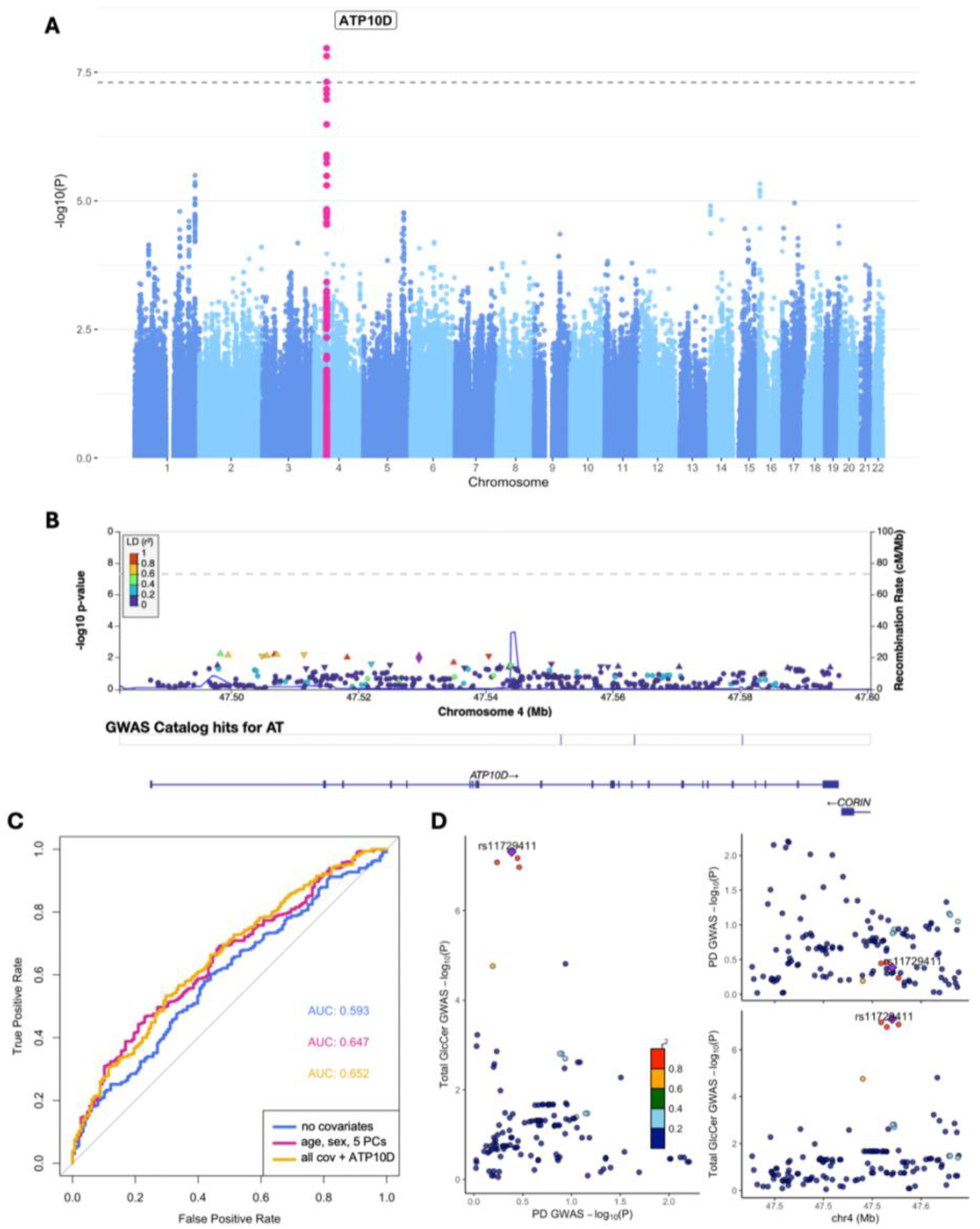
**A)** Manhattan plot of log adjusted p-values at each genomic position for GWAS of total GlcCer levels in the PPMI cohort adjusted for age, sex, disease status, and 5 PCs, **B)** Locuszoom Manhattan plot of the *ATP10D* gene region in the PD GWAS, **C)** Area under the curve analysis for logistic regressions of total GlcCer with PD status, with and without adjustment for the lead *ATP10D* GWAS hit, **D)** Colocalization plot comparing *ATP10D* locus p-values in the total GlcCer levels GWAS and the PD GWAS.

In plasma GlcCer isoforms, the *ATP10D* locus was associated with total GlcCer and with the C24:1 isoform, while almost reaching significance and exhibiting a consistent direction of effect with other GlcCer isoforms (C16, C20, C22, C23, and C24, Figure 1A, Table 1, Supplementary Figure 2). Many of the lead variants across the isoforms were in strong linkage disequilibrium (LD), supporting a common haplotype/variant driving the effect (Supplementary Figure 3). Interestingly, the effects of ATP10D on GlcCer levels were much stronger than GBA1 variants associated with PD. While rs6827604 in the *ATP10D* locus had a very strong effect on GlcCer levels (beta=0.41, SE=0.070, p=1.076e-08, Table 1), the three most common *GBA1* variants associated with PD had almost no effect on GlcCer (p.E326K; beta=0.067, SE=0.26, p=0.79, p.T369M: beta=-0.071, SE=0.34, p=0.83, p.N370S: beta=-0.37, se=0.40, p= 0.36), indicating that *ATP10D* is a much stronger regulator of plasma GlcCer than *GBA1*. We further investigated the *ATP10D* locus in the largest PD GWAS summary statistics^14^ to determine if *ATP10D* plays a role in PD risk, and no variants were associated with PD after Bonferroni correction (Figure 1B, Supplementary Table 4).

**Table 1.** Lead *ATP10D* SNPs for plasma GlcCer isoforms from GWASs in PPMI.

| GlcCer Isoform | Lead SNP | Beta | SE | P-value |
| --- | --- | --- | --- | --- |
| C16 | rs11729411 | 0.034 | 0.0062 | 8.83e-08 |
| C18 | rs4694857 | 0.0043 | 0.00098 | 1.35e-05 |
| C20 | rs6827604 | 0.0068 | 0.0013 | 3.831e-07 |
| C22 | rs11729411 | 0.067 | 0.013 | 2.14e-07 |
| C23 | rs6827604 | 0.039 | 0.0077 | 7.804e-07 |
| C24:1 | rs6827604 | 0.12 | 0.020 | 1.117e-08 |
| C24 | rs6827604 | 0.16 | 0.030 | 9.803e-08 |
| Total | rs6827604 | 0.41 | 0.070 | 1.076e-08 |

GlcCer, glucosylceramide; SNP, single nucleotide polymorphism; GWAS, genome-wide association study; PPMI, Parkinson's Progression Markers Initiative; SE, standard error.

Colocalization analysis of the *ATP10D* SNPs from the total GlcCer GWAS and PD GWAS demonstrated evidence for one unique causal signal for GlcCer levels (PPH1 = 95.4%) and no evidence of colocalization (PPH4 < 0.8%), supporting that the *ATP10D* locus, and importantly, plasma GlcCer levels, are not causal for PD (Figure 1D). The positive association of *ATP10D* and GlcCer levels was replicated in a GWAS of the Centogene cohort that included C16, C18, C22, C24:1, and C24 isoforms (Supplementary Table 5). All top associations in the Centogene GWAS were in strong LD with the top associations in PPMI, again suggesting one associated haplotype or variant that drives the association (Supplementary Figure 3).

We then aimed to examine whether GlcCer levels are associated with PD. We performed logistic regressions of the plasma GlcCer isoforms and PD status with *GBA1* p.N370S and *LRRK2* p.G2019S carriers removed, and found that all isoforms were associated with PD risk (Table 2). We then performed receiver operating characteristic (ROC) area under the curve (AUC) analysis, showing that GlcCer is not a good predictor of PD status (Figure 1C). Adjusting for *ATP10D* SNPs associated with GlcCer levels did not improve the predictive ability of the model.

**Table 2.** Logistic regression for plasma GlcCer isoforms with PD status in PPMI adjusted for age, sex, and 5 PCs.

| GlcCer Isoform | GlcCer x PD |  |  |
| --- | --- | --- | --- |
|  | Beta | SE | P-value |
| C16 | 3.99 | 1.29 | 0.002 |
| C18 | 23.2 | 7.03 | 0.00098 |
| C20 | 19.8 | 5.2 | 0.00014 |
| C22 | 1.96 | 0.64 | 0.0024 |
| C23 | 2.27 | 0.86 | 0.0083 |
| C24 | 0.74 | 0.23 | 0.001 |
| C24:1 | 1.2 | 0.33 | 0.00033 |
| Total | 0.33 | 0.095 | 0.00059 |

GlcCer, glucosylceramide; PC, principal component; SNP, single nucleotide polymorphism; PD, Parkinson's disease; PPMI, Parkinson's Progression Markers Initiative; SE, standard error.

## Discussion

Our study shows that the strongest modifier of plasma GlcCer levels is *ATP10D*. Since variants in *ATP10D* that have a strong effect on GlcCer levels are not associated with PD (Figure 1D), it is unlikely that GlcCer levels have a role in the pathogenesis of PD. This observation is further supported by our colocalization analysis. Previous studies have shown that GlcCer levels are statistically different in PD versus controls,^15^ as we have replicated here. Therefore, it seems that while GlcCer levels are somewhat different on average between PD patients and controls, this association is not causative, and also cannot be used as a stand-alone biomarker to predict PD status, as demonstrated in our ROC-AUC analysis.

*ATP10D* encodes ATPase Phospholipid Transporting 10D, which is located mainly on the endoplasmic reticulum and plasma membranes. This protein is a flippase whose main function is to flip glucosylceramides across membranes,^16^ and it has already been shown that *ATP10D* variants affect GlcCer levels,^17^ as we have demonstrated here. Recently, a clinical trial (MOVES-PD, clinicaltrials.gov ID: NCT02906020) targeting GlcCer by inhibiting the enzyme producing it, glucosylceramide synthase, had been completed.^9^ The drug tested in this trial, venglustat, demonstrated excellent target engagement, as GlcCer levels were significantly reduced as expected both in plasma and CSF. However, there was no clinical benefit for the participants who received venglustat, who in fact did worse compared to those who received placebo. Our findings suggesting that GlcCer levels are likely to have no pathogenic role in PD, combined with the findings of the MOVES-PD trial, may suggest that altering GlcCer levels is not a good therapeutic target for *GBA1*-PD.

Our study has several limitations. Our analysis is focused on plasma GlcCer and not brain GlcCer levels, which is not available. CSF GlcCer levels may serve as surrogate data for brain GlcCer levels, however the data from the CSF from PPMI was less robust. Despite this, CSF GlcCer levels had good correlation with plasma GlcCer levels (r=0.32, p=2.4×10^−15^), which allowed us to use plasma GlcCer levels. Despite the small sample size, the results showing that *ATP10D* is the strongest regulator of GlcCer levels were confirmed in the Centogene data as well as in a previous report.

To conclude, our results argue against targeting GlcCer in *GBA1*-PD, and suggest that better understanding of the mechanism underlying *GBA1*-PD may lead to better therapeutic targets.

## Data Availability

All code for analyses used in this project can be found at https://github.com/gan-orlab/GlcCer_GWAS/. PPMI data used in the present study can be accessed by qualified researchers through completion of a data access application (https://www.ppmi-info.org/access-data-specimens/download-data). Data from the Centogene cohort may be made available on request due to restrictions. All PPMI plasma GlcCer GWAS summary statistics are publicly available in the GWAS catalog (https://www.ebi.ac.uk/gwas/home).

## Acknowledgements

Data used in the preparation of this article were obtained from the Parkinson’s Progression Markers Initiative (PPMI) database (www.ppmi-info.org/access-data-specimens/download-data). For up-to-date information on the study, visit www.ppmi-info.org. PPMI – a public-private partnership – is funded by the Michael J. Fox Foundation for Parkinson’s Research and funding partners, including 4D Pharma, Abbvie, AcureX, Allergan, Amathus Therapeutics, Aligning Science Across Parkinson’s, AskBio, Avid Radiopharmaceuticals, BIAL, BioArctic, Biogen, Biohaven, BioLegend, BlueRock Therapeutics, Bristol-Myers Squibb, Calico Labs, Capsida Biotherapeutics, Celgene, Cerevel Therapeutics, Coave Therapeutics, DaCapo Brainscience, Denali, Edmond J. Safra Foundation, Eli Lilly, Gain Therapeutics, GE HealthCare, Genentech, GSK, Golub Capital, Handl Therapeutics, Insitro, Jazz Pharmaceuticals, Johnson & Johnson Innovative Medicine, Lundbeck, Merck, Meso Scale Discovery, Mission Therapeutics, Neurocrine Biosciences, Neuron23, Neuropore, Pfizer, Piramal, Prevail Therapeutics, Roche, Sanofi, Servier, Sun Pharma Advanced Research Company, Takeda, Teva, UCB, Vanqua Bio, Verily, Voyager. We would like to thank the research participants for contributing to this study. We thank Meron Teferra for her assistance. KS is supported by a post-doctoral fellowship from the Canada First Research Excellence Fund (CFREF), awarded to McGill University for the Healthy Brains for Healthy Lives initiative (HBHL). Z.G.O. is supported by the Fonds de recherche du Québec— Santé (FRQS) Chercheurs-boursiers award and is a William Dawson Scholar. E.N.S is supported by a graduate student award from Parkinson Canada. This study was financially supported through grants from the Galen and Hilary Weston Foundation and the Michael J. Fox Foundation (MJFF). This work was supported in part by the Intramural Research Program of the National Institute on Aging (NIA), and the Center for Alzheimer’s and Related Dementias (CARD), within the Intramural Research Program of the National Institute on Aging and the National Institute of Neurological Disorders and Stroke (1ZIA-AG000546).

## Author Contributions

E.N.S., M.T, H.I, and Z.G.O. contributed to the conception and design of the study. All authors contributed to the acquisition and analysis of data. E.N.S., A.J. and Z.G.O. contributed to drafting a significant portion of the manuscript or figures.

