## Supplementary Text for "Plasma glucosylceramide levels are regulated by *ATP10D* and are not involved in Parkinson’s disease pathogenesis"

**Parkinson’s Progression Markers Initiative plasma glucosylceramide extraction and analysis.**

Glucosylceramide (GlcCer) isoforms in plasma were extracted with protein precipitation in extraction solution (acetonitrile/methanol/formic acid, 50/50/0.2, v/v/v, with 5 mM ammonium formate). An internal standards cocktail of 1.34 ng/mL of C12:0 GlcCer was used in the extraction solvent. The mixtures were vortexed, centrifuged, and final supernatant was transferred to 384-well plates for quantitative GlcCer analysis using liquid chromatography with tandem mass spectrometry (LC-MS/MS). GlcCer was analyzed with a Waters 2.1 x 100 mm Cortecs HILIC column at a flow rate of 0.5 mL/min and a Waters Acquity binary solvent system using mobile phase A and mobile phase B (79/20/1, methanol/water/acetic acid, v/v/v, with 5 mM ammonium acetate). LC eluents were analyzed by a triple quadrupole mass spectrometer in MRM mode, and GlcCer standards were used to generate calibration curves. Further details can be accessed by approved researchers through the PPMI website under Project 135.

**Centogene cohort methodology.**

892 PD patients and 139 healthy controls, including both GBA carriers and noncarriers, for the Centogene cohort were recruited with the ROPAD study, which is an observational clinical study in a multicenter international setting.^1^ Genomic DNA was extracted from dried blood spots (DBS) and sequenced using the Illumina high throughput sequencing technology according to manufacturer’s protocol (Illumina Inc) at Centogene. Sample-level quality control included kinship analysis to remove related individuals and sex imputation from genetic data, with undetermined cases documented. Variant-level filtering included removal of SNPs with poor quality, low call rates, deviations from Hardy-Weinberg equilibrium, and rare minor allele frequencies (MAF). Additional filtering included read depth (>10), genotype quality (GQ>20), variant quality (QUAL>4). Linear regression was used to assess the association between genetic variants and phenotypes. Covariates were automatically selected using component-wise ANOVA and Wald tests to control for confounding factors.

Biomaterial was obtained from DBS from EDTA whole human blood dropped on CentoCard® (Centogene, Rostock, Germany) filter paper. The pure analytical standards were purchased through German suppliers from Matreya LLC (State College, Pennsylvania, USA) and Avanty Polar Lipids Inc (Birmingham, Alabama, USA). All standards were purchased in high purity with the exception of the glucosyl(β)Ceramides (d18:1/22:0) and (d18:1/24:0). The latter two were purchased as a mixture from buttermilk (27% and 23%respectivelly). From each filter card, five 3.2 mm disks were cut with a puncher (PerkinElmer, Germany) representing the equivalent of 15.5 µL blood. The paper disks were mixed with an extraction solution containing the internal standard (Lyso-Gb2) in a DMSO:water:ethanol, 3:2:10 (v:v:v) mixture and incubated for one hour at 37 °C and 700 rpm. The solution was transferred onto an PTFE (AcroPrep) filter plate and filtered by centrifugation for 5 min at 3500 rpm to remove paper debris, and afterwards loaded into the autosampler. The samples were measured using an LC-MRM-MS method on an Acquity UPLC (Waters GmbH, Eschborn, Germany) coupled with an TQ6500 mass spectrometer (AB-Sciex, Darmstadt, Germany). An ACE 3 C8 50x2.1mm UPLC column (MZ-Analysentechnik GmbH, Mainz, Germany) was used to separate the analytes. A mixture of Acetone:Acetonitrle (1:1, v:v) containing 50 mM formic acid was used as organic phase and 50 mM formic acid in LC-MS grade water was used as aqueous phase. A six-minute gradient from 40% to 100% organic phase was used to separate the analytes before quantifying them by MS under a flow rate of 0.89 mL/min. and a column temperature of 60°C. The MS analysis was performed in MRM mode: Ion spray voltage of 5500 V, Source temperature of 500 °C, curtain gas 40 psi and a collision energy of 55 V for the Ceramides and 35 V for the Sphingosines. Monitored MRM transitions were 700.57 > 264.27 for GlcCer 16:0, 728.6 > 264.27 for GlcCer 18:0, 784.66 > 264.27 for GlcCer 22:0, 810.68 > 264.27 for GlcCer 24:1, 812.69 > 264.27 for GlcCer 24:0, 462.3 > 282.2 for lyso-Gb1 and 624.3 > 282.2 for the lyso-Gb2. The latter was used as internal standard. This LC/MS method does not differentiate between the glucosyl- and galactosyl- isomers of the analytes. However, the galactosyl isomer concentrations are low relative to the glucosyl isomers and thus negligible as was shown for the glucosyl-/galactosylsphingosine isomer pair in human plasma.^2^ Hence, throughout this work, the measured concentrations are attributed to the glucosyl isomers. An 11-points calibration line with glucosylceramides and Lyso-Gb1 concentrations between 0 and 200 ng/mL and a linear fitting regression through 0 was used to quantify the biological samples. Per calibration point, 15.5 µL of solution containing each analyte was processed and measured in the same way as the equivalent to 15.5 µL blood extract from the DBS samples. Analysis and quantification were performed by using the Analyst 1.6.2 software (AB Sciex Germany GmbH, Darmstadt, Germany).
