## Supplementary Data for "Plasma glucosylceramide levels are regulated by *ATP10D* and are not involved in Parkinson’s disease pathogenesis"

**Supplementary Figures.**

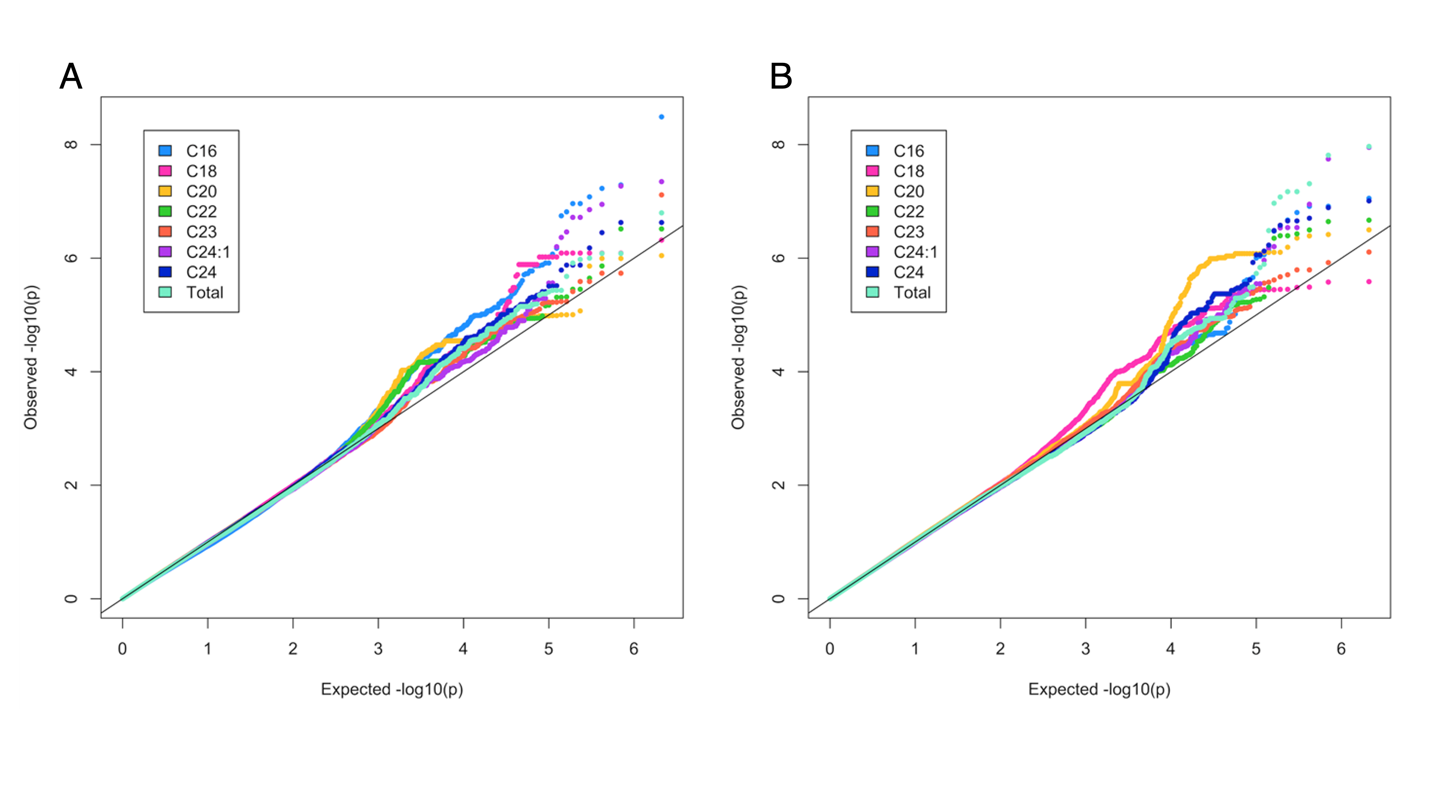

**Supplementary Figure 1**. QQ plots of all Plasma GlcCer isoform GWAS p-values in PPMI.

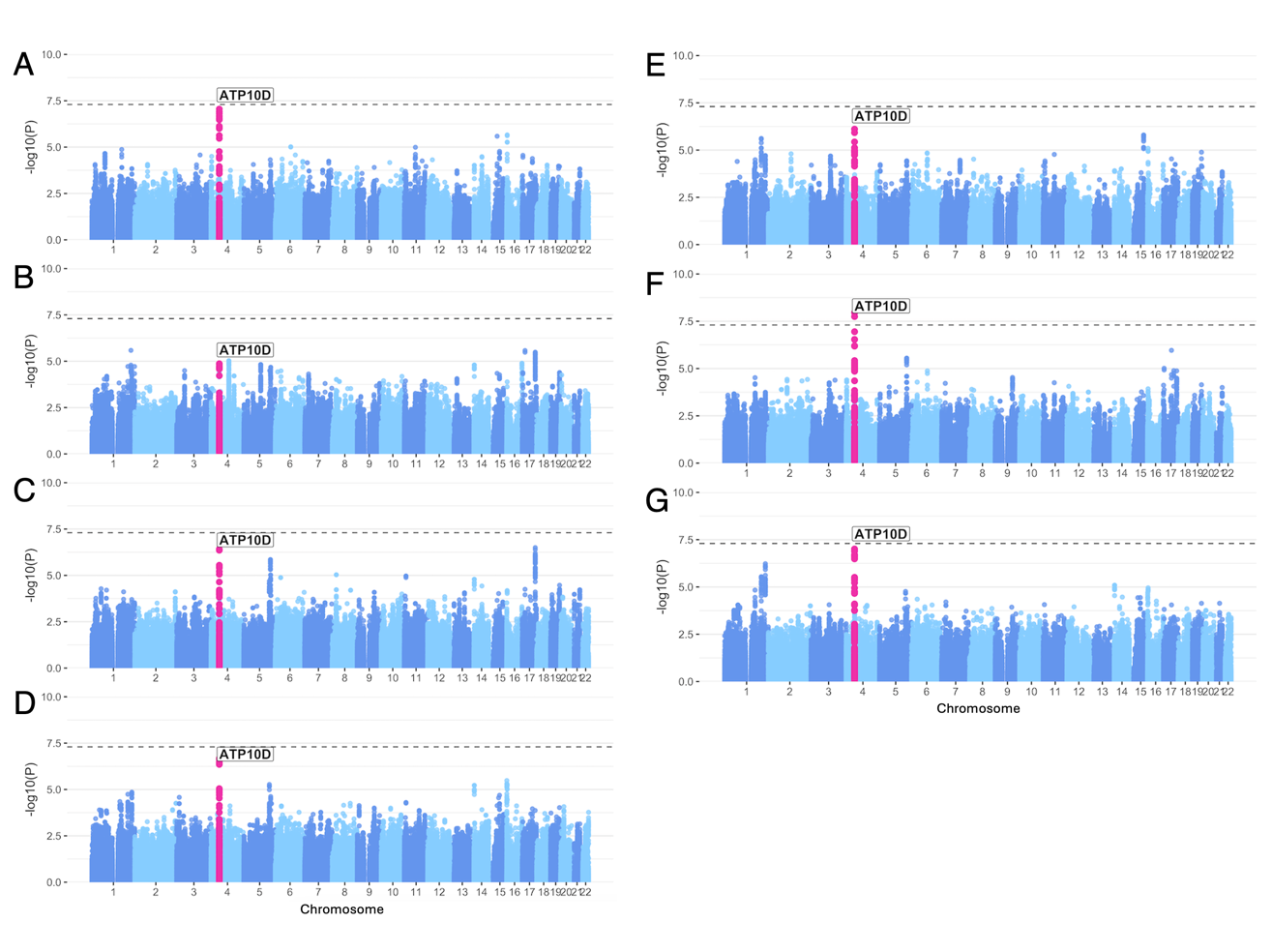

**Supplementary Figure 2.** Manhattan plots of genome-wide association studies of plasma GlcCer levels in the PPMI cohort for A) C16, B) C18, C) C20, D) C22, E) C23, F) C24:1, and H) total isoforms adjusted for age, sex, disease status, and 5 PCs.

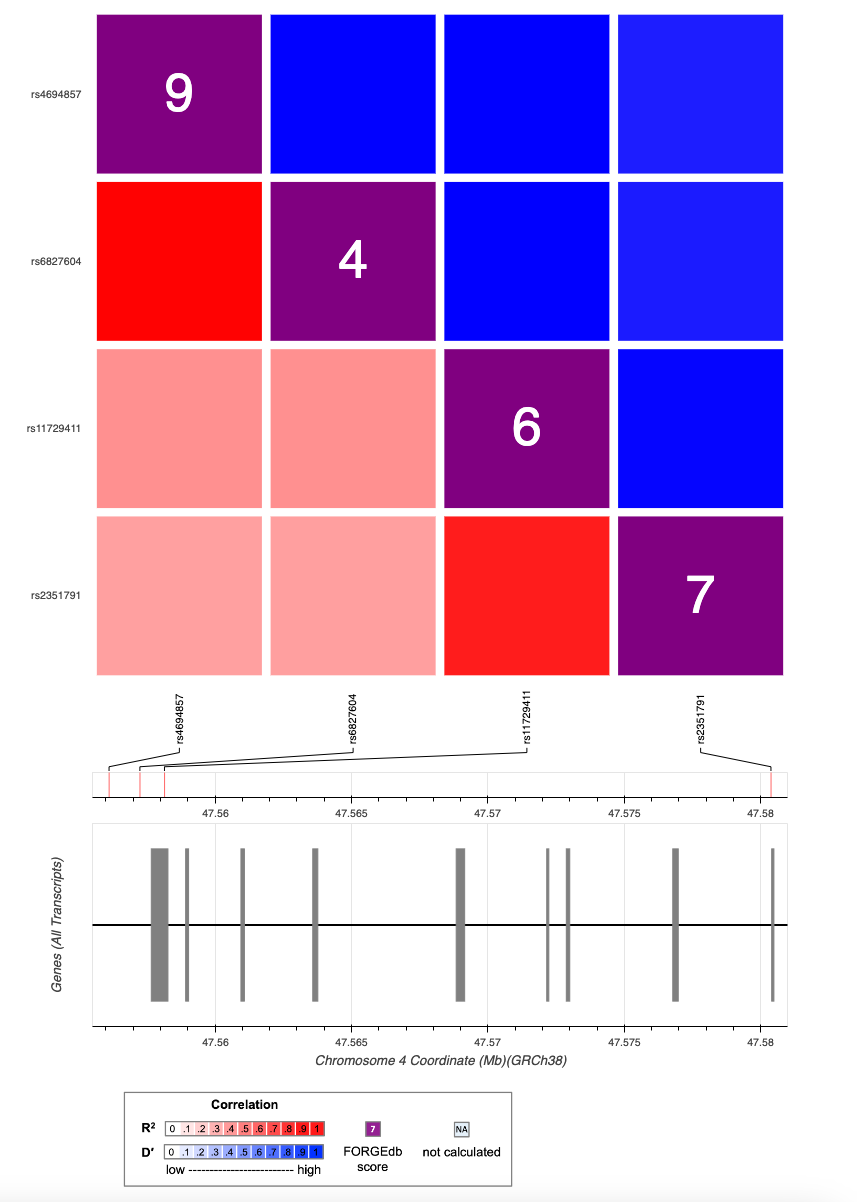

**Supplementary Figure 3.** LDlink linkage disequilibrium heatmap depicting R^2^ and D′ for lead *ATP10D* SNPs from plasma GlcCer GWASs.

**Supplementary Tables.**

**Supplementary Table 1.** Cohort demographics for PPMI participants with available genotype data.

| **Disease Group** | **Males (%)** | **Females (%)** | **Mean Age (±SD)** |
| --- | --- | --- | --- |
| Parkinson’s Disease | 66.9 | 33.1 | 61.9 (9.6) |
| Healthy Controls | 65.9 | 34.1 | 61.1 (10.7) |
| SWEDD | 64.7 | 35.3 | 60.9 (9.8) |

PPMI, Parkinson’s Progression Markers Initiative; SD, standard deviation; SWEDD, scan without evidence of dopaminergic deficit.

**Supplementary Table 2.** Mean glucosylceramide levels for PPMI participants following quality control.

| **Tissue** | **Isoform** | **Median** | **SD** |
| --- | --- | --- | --- |
| CSF | C16 | 0.27 | 0.2 |
|  | C18 | 0.62 | 0.24 |
|  | C20 | 0.054 | 0.11 |
|  | C22 | 0.6 | 0.5 |
|  | C23 | 0.46 | 0.38 |
|  | C24-1 | 2.24 | 1.78 |
|  | C24 | 2.15 | 1.43 |
|  | Total | 6.37 | 4.35 |
| Plasma | C16 | 0.32 | 0.085 |
|  | C18 | 0.053 | 0.017 |
|  | C20 | 0.075 | 0.022 |
|  | C22 | 0.58 | 0.17 |
|  | C23 | 0.41 | 0.13 |
|  | C24-1 | 1.055 | 0.34 |
|  | C24 | 1.69 | 0.49 |
|  | Total | 4.067 | 1.17 |

PPMI, Parkinson’s Progression Markers Initiative; SD, standard deviation; CSF, cerebrospinal fluid

**Supplementary Table 3.**  Lambdas for CSF and Plasma GlcCer GWASs in PPMI.

| **Tissue** | **Isoform** | **Lambda** |
| --- | --- | --- |
| CSF | C16 | 0.97 |
|  | C18 | 0.99 |
|  | C20 | 1.015 |
|  | C22 | 0.98 |
|  | C23 | 1.0 |
|  | C24-1 | 0.99 |
|  | C24 | 0.99 |
|  | Total | 0.98 |
| Plasma | C16 | 1.015 |
|  | C18 | 1.026 |
|  | C20 | 1.029 |
|  | C22 | 1.01 |
|  | C23 | 1.0089 |
|  | C24-1 | 1.0 |
|  | C24 | 1.00047 |
|  | Total | 1.0033 |

CSF, cerebrospinal fluid; GlcCer, glucosylceramide; GWAS, genome-wide association study.

**Supplementary Table 4.** ATP10D Variants below nominal significance in Nalls et al. 2019 PD risk GWAS.

| **SNP** | **rsID** | **Beta** | **SE** | **P-value** |
| --- | --- | --- | --- | --- |
| 4:47498294 | rs4393976 | 0.0342 | 0.0124 | 0.005688 |
| 4:47506788 | rs7692584 | 0.0346 | 0.0126 | 0.006234 |
| 4:47511448 | rs12501399 | -0.0343 | 0.0126 | 0.006332 |
| 4:47507193 | rs7699242 | 0.0343 | 0.0126 | 0.006467 |
| 4:47499506 | rs5026454 | 0.0337 | 0.0125 | 0.007036 |
| 4:47505636 | rs62300111 | 0.0331 | 0.0124 | 0.00767 |
| 4:47504833 | rs12640836 | -0.0329 | 0.0124 | 0.007754 |
| 4:47540509 | rs62297979 | -0.0342 | 0.0129 | 0.007839 |
| 4:47518239 | rs62297974 | 0.0327 | 0.0126 | 0.009621 |
| 4:47529525 | rs12512121 | 0.0327 | 0.0127 | 0.009827 |
| 4:47534994 | rs62297978 | 0.0314 | 0.0135 | 0.02027 |
| 4:47522029 | rs4499679 | -0.026 | 0.0116 | 0.02495 |
| 4:47550356 | rs139377928 | -0.1496 | 0.0668 | 0.02511 |
| 4:47524961 | rs12509973 | -0.0272 | 0.0124 | 0.02859 |
| 4:47497848 | rs112494819 | 0.0472 | 0.0219 | 0.03114 |
| 4:47544012 | rs4694853 | -0.0218 | 0.0102 | 0.03357 |
| 4:47543891 | rs4694852 | 0.0216 | 0.0102 | 0.03485 |
| 4:47560870 | rs140919542 | 0.1344 | 0.0643 | 0.03652 |
| 4:47558132 | rs4694857 | -0.02 | 0.0095 | 0.03659 |
| 4:47542271 | rs12651326 | 0.0211 | 0.0102 | 0.03763 |
| 4:47559262 | rs6827604 | -0.0198 | 0.0095 | 0.03807 |
| 4:47570454 | . | 0.023 | 0.0111 | 0.03861 |
| 4:47514685 | rs33995001 | -0.0195 | 0.0095 | 0.03973 |
| 4:47543657 | . | 0.0207 | 0.0101 | 0.0413 |
| 4:47594204 | . | 0.0347 | 0.017 | 0.04138 |
| 4:47587550 | . | 0.0342 | 0.017 | 0.04414 |
| 4:47536695 | rs28493195 | -0.0203 | 0.0101 | 0.04449 |
| 4:47542429 | rs4283632 | -0.0206 | 0.0103 | 0.04469 |
| 4:47542160 | . | -0.0203 | 0.0101 | 0.04504 |
| 4:47587877 | rs11932436 | -0.034 | 0.0171 | 0.04693 |
| 4:47544639 | . | -0.028 | 0.0142 | 0.04781 |
| 4:47511781 | . | -0.0192 | 0.0097 | 0.04908 |

PD, Parkinson’s disease; GWAS, genome-wide association study; SNP, single nucleotide polymorphism; SE, standard error.

**Supplementary Table 5.** Lead *ATP10D* SNPs for plasma GlcCer isoforms from GWASs in the Centogene cohort.

| **GlcCer Isoform** | **Lead SNP** | **Beta** | **SE** | **P-value** |
| --- | --- | --- | --- | --- |
| C16 | rs2351791 | 33.74 | 7.03 | 1.82e-06 |
| C18 | rs2351791 | 2.93 | 0.66 | 9.40e-06 |
| C22 | rs2351791 | 56.42 | 8.34 | 2.26e-11 |
| C24:1 | rs2351791 | 16.13 | 4.55 | 0.0004 |
| C24 | rs2351791 | 101.1 | 15.94 | 3.38e-10 |

GlcCer, glucosylceramide; SNP, single nucleotide polymorphism; GWAS, genome-wide association study; PPMI, Parkinson’s Progression Markers Initiative; SE, standard error.
